# Sensitivity of UK Covid-19 deaths to the timing of suppression measures and their relaxation

**DOI:** 10.1101/2020.05.09.20096859

**Authors:** John Dagpunar

**Affiliations:** University of Southampton, UK

**Keywords:** Covid-19, SEIR model, lockdown, suppression, halving time, doubling time

## Abstract

In this paper I examine the sensitivity of total UK Covid-19 deaths and the demand for intensive care and ward beds, to the timing and duration of suppression periods during a 500 day period. This is achieved via a SEIR model. Using an expected latent period of 4.5 days and infectious period of 3.8 days, R_0_ was first estimated as 3.18 using observed death rates under unmitigated spread and then under the effects of the total lockdown (R_0_ =0.60) of 23 March. The case fatality rate given infection is taken as 1%. Parameter values for mean length of stay and conditional probability of death for ICU and non-ICU hospital admissions are guided by Ferguson et al.(2020). Under unmitigated spread the model predicts around 600,000 deaths in the UK. Starting with one exposed person at time zero and a suppression consistent with an R_0_ of 0.60 on day 72, the model predicts around 39,000 deaths for a first wave, but this reduces to around 11,000 if the intervention takes place one week earlier. If the initial suppression were in place until day 200 and then relaxed to an R_0_ of 1.5 between days 200 and 300, to be followed by a return to an R_0_ of 0.60, the model predicts around 43,000 deaths. This would increase to around 64,000 if the release from the first suppression takes place 20 days earlier. The results indicate the extreme sensitivity to timing and the consequences of even small delays to suppression and premature relaxation of such measures.

## 1. The SEIR model

In order to model the spread of total Covid-19 deaths in the UK over the next 500 days say, one can build a detailed stochastic microsimulation model that includes spatial and age heterogeneity as in Davies et al. (2020) and Ferguson et al. (2020). In order to gain some understanding of the sensitivity with respect to the timing of suppression measures and their relaxation, a simple deterministic SEIR model, see for example Hethcote (2000) and Blackwood and Childs (2008), can be useful. That is the approach used in this paper. SEIR is a compartment based model with transitions as shown in Diagram 1. *S* stands for the number of susceptibles, those who have not yet been infected. Initially, almost the entire population is in that category. *E* is the number who are currently exposed, that is they are infected but not yet infectious. After a latent period, an exposed person becomes infectious and is able to infect susceptible persons. *I* is the number who are currently infectious. Initially, both *I* and *E* are small numbers in comparison to *N*, the population size. Each infectious person remains so for an infectious period. An infective person is one who is either exposed or infectious. Finally, *R* stands for those who are removed in the sense they have either recovered or died. In the simple form of the model considered here it is assumed that *S + E + I + R = N*. The SEIR model is more appropriate than a SIR model for Covid-19 as SIR does not separate the exposed and infectious states.

A key controllable parameter is the reproduction number *R*_0_ which is the expected number of susceptibles that an infector will infect, when the susceptible proportion is close to 1. It is thought that a significant proportion of those infected may be asymptomatic or pre-symptomatic in which case they might spend all or part of the infectious period unknowingly infecting susceptibles, although there is a question as to how much virus they shed. On the other hand, those who are symptomatic may spend some of that time in self-isolation which reduces the *R*_0_ for them. One of the simplifying assumptions of the model developed here is that people can transmit the virus throughout their infectious period.

A person will transition between these compartments. The SEIR model will keep track of how many are in each compartment by noting the rate at which people join and leave a compartment. The key dynamic element of this process is the transmission rate *β* at which an infectious person infects susceptibles.

**DIAGRAM 1.**
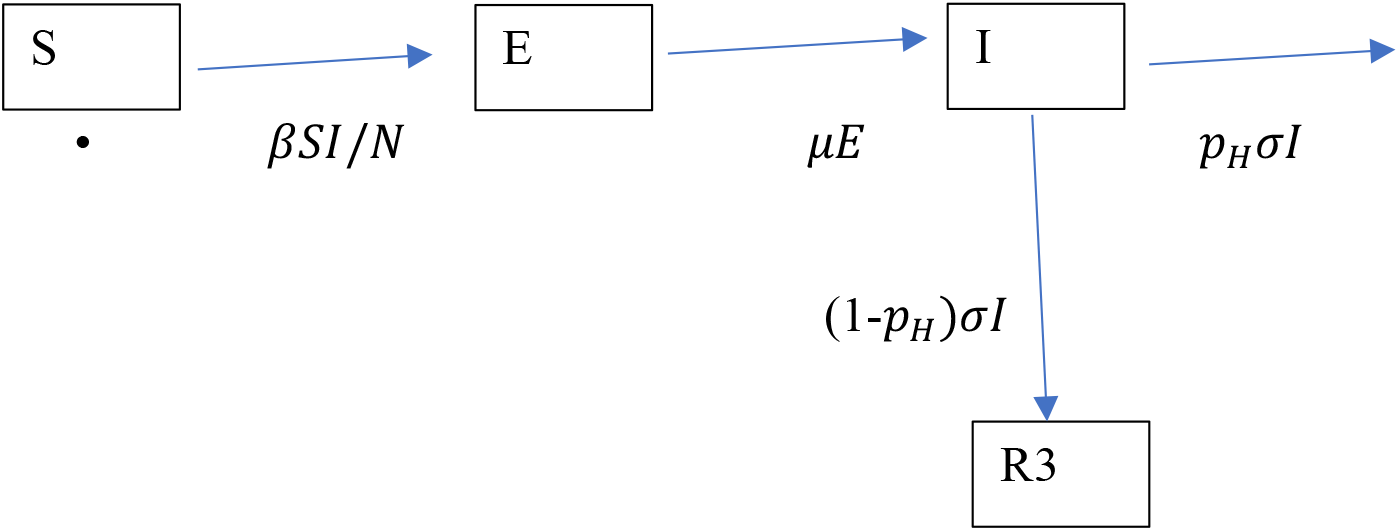

Diagram 1 shows the rates of movement between the compartments. The mean latent and infectious periods are respectively 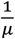 and 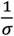, while *p_H_* is the probability that an infectious person is admitted to hospital. R3 is the cumulative number who do not require hospital treatment. It is assumed that all such persons survive, but it may be more accurate to say with hindsight that it may include an appreciable number of people in the community who never went to hospital and did not survive. In that sense the model underestimates the cumulative number of deaths. The model is an idealisation of what happens in practice as some people will enter hospital while still infectious and some will self-isolate during the infectious state. This can be compensated for by a suitable choice of *σ*.

## 2. Bed Occupancy

Apart from predicting the number of deaths it is also important to predict the demand for ward and intensive care beds. The UK National Health Service (NHS) has approximately 150,000 ward beds and around 4,200 Intensive care beds. Clearly, one could not expect to access all of these as the occupancy rate is high in normal times. There is the option of building surge capacity. Ideally, that would need to be matched by similar increases in medical, nursing, and ancillary staff; equipment such as ventilators; personal protective equipment; and testing for presence of the virus, all of which have become major issues. Turning now to what happens in hospitals, Diagram 2 gives a schematic view with the transition rates.

**DIAGRAM 2.**
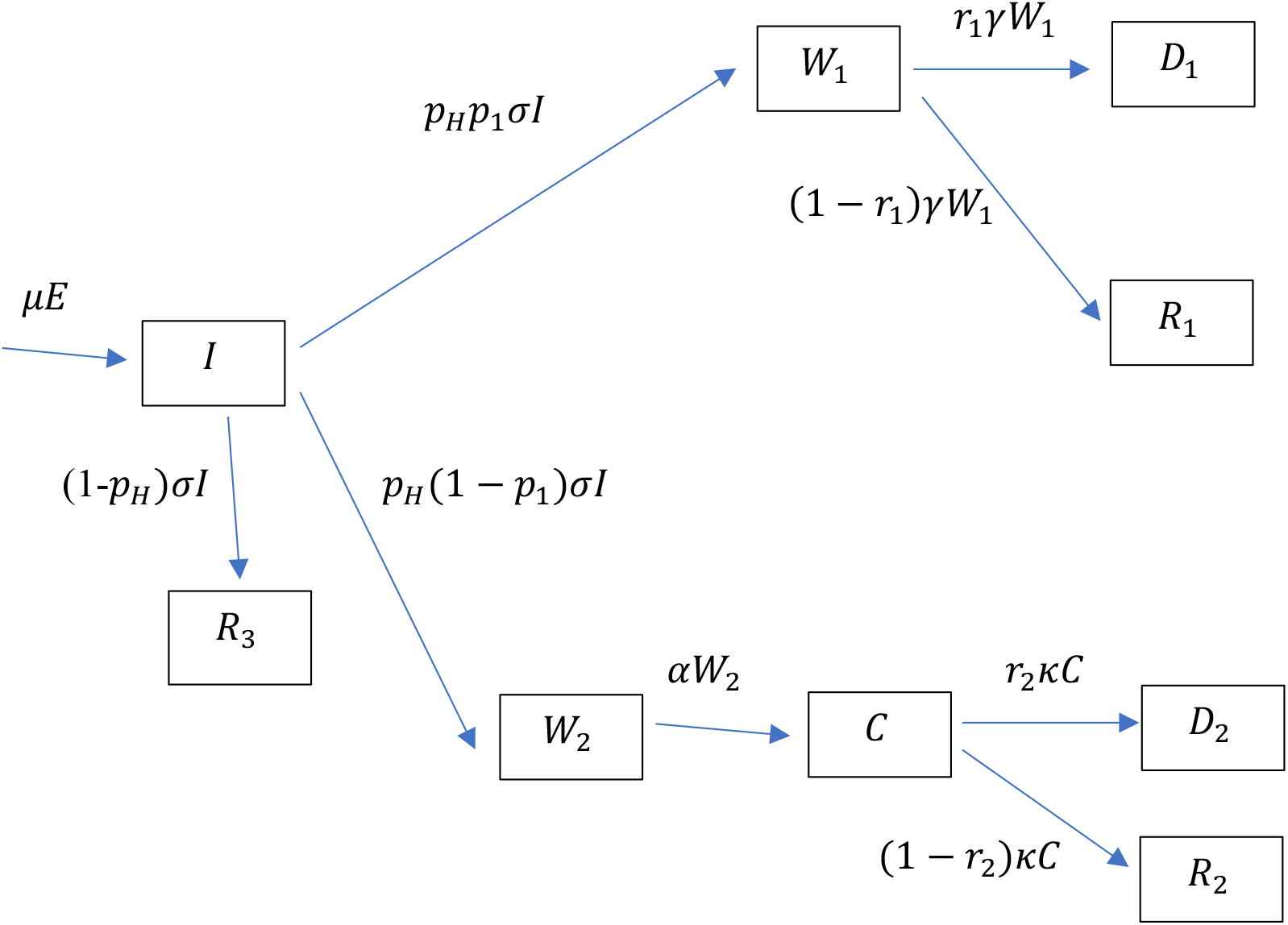

An infectious person will move with probability *p_H_* to hospital. Of these a proportion *p*_1_,will never require an intensive care bed and will occupy a ward bed. Of those a proportion *r*_1_ will die and the rest will recover. Either way their average length of stay will be 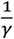 days. The remaining proportion 1 – *p*_1_ will spend an average of 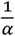 days in a ward bed followed by an average of 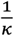 days in intensive care. A proportion *r*_2_ of those entering intensive care will die. For simplicity, those who recover are not re-directed to a ward bed. The symbol in each compartment gives the number of persons currently occupying it, *D*_1_ and *D*_2_ being the cumulative number of deaths from non-ICU and ICU patients respectively.

## 3. The Dynamics

Diagrams 1 and 2 lead directly to differential equations (1-8)

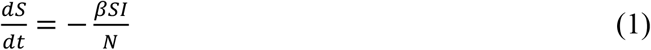

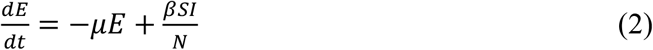

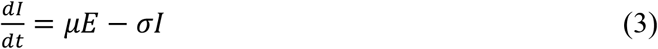

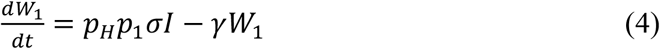

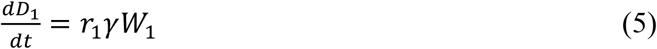

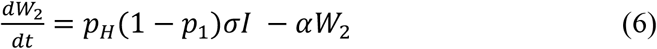

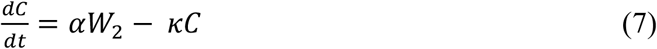

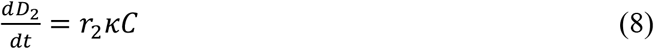

## 4. Epidemic growth and decline rate in early stages

In this section we obtain the important results for the case when *S/N ≈* 1. Equations (2-3) can be written as

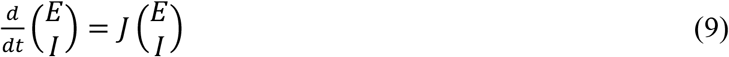

where 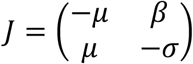. The eigenvalues of *J are*

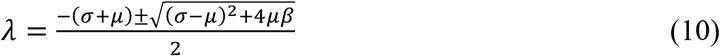

The negative root will give a solution that decays rapidly and can be ignored, see for example Ma (2020). So, while the susceptible proportion is close to 1 we have the solution

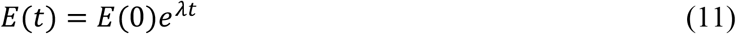

and

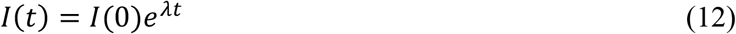

where

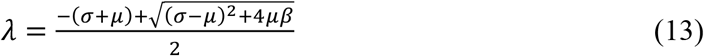

that is

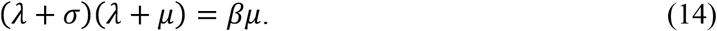

The expected number of secondary infections from one infectious person is

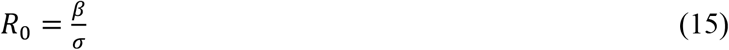

For the special case of *σ = μ*

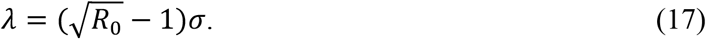

Substituting (10) into (9)

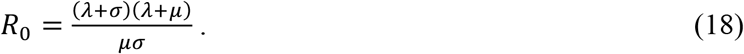

While *λ >* 0 the doubling time is

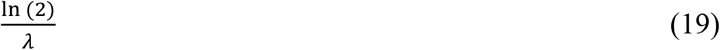

and when *λ < 0* the half-life (halving time) is

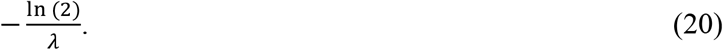

## 5. Estimation of *R*_0_

From (11) and (12), in the early stages of the epidemic, current infectious numbers *I*(*t*), new daily infectious numbers *μE*(*t*), and daily deaths 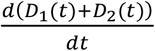, will all^1^ change at rate *λ* so one could use any of these to estimate *λ*. In the UK there was a large proportion^2^ of infectious persons (including asymptomatic and pre-symptomatic) who were never tested and the availability of tests changed markedly over time. Therefore, it is felt that the number of positive test results is not a reliable proxy for the number of infectious persons.

For this reason, *λ* was estimated by fitting the logarithm of daily deaths non-parametrically in the period 11 March to 2 April. The dates were selected as to be largely uninfluenced by the gentle suppression introduced on 16 March and the full lockdown beginning 23 March, given that there is a lag between exposure and death of the order of 24 days. The fit gave *λ* = 0.189, 95% confidence interval (0.172, 0.206) with the amount explained by the regression as *R*^2^ = 0.96. The estimated doubling time in the early stages of unmitigated spread is 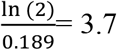 days.

In order to estimate *R*_0_ using (18) we need not only *λ*, but estimates of mean latent and infectious periods. Ferguson et al. (2020) assumed an incubation period of 5.1 days with a 12 hour pre-symptomatic infectious period for some, together with a 6.5 day mean generation interval. Anderson et al. (2020) thought the incubation period to be around 5-6 days with a serial interval of similar duration. Davies et al. (2020) assumed a gamma distributed latent period with mean and standard deviation of 4 and 2 days respectively, a mean incubation period of 5.5 days, and a serial interval of 6.5 days. In this paper we take the mean latent and infectious period to be 4.5 and 3.8 days respectively, leading to a crude estimate of the serial interval to be 4.5+0.5×3.8 = 6.4 days. Using these in (18) gives *R*_0_ *=* 3.18. The death data was that pertaining to English hospital deaths by date of death.^3^ This data set was chosen as many other accessible sets are by date of recording the death which can be several days after actual death. We assumed that the growth/decline rate *λ* is similar in all four countries of the UK. The model assumes all deaths occur in hospital and applies to the entire UK of population 67 million. For illustrative purposes this is considered reasonable although in the event many deaths occurred in the community including care homes. The model calculates bed occupancy assuming an ideal scenario where all who need hospital care receive it.

It is assumed that 1% of those infected die and 4.4% of those infected require hospital treatment. Of these 70% will never require intensive care and spend an average of 8 days in a ward bed. Of these 11% will die. Of the remaining 30%, all will spend 6 days in a ward followed by 10 days in intensive care and 50% of these will die, These parameter values give a case fatality rate given infection of 1%. These figures are adapted from those of Ferguson et al. (2020).

## 6. Unmitigated Spread

Table 1 summarises the model results over a period of 500 days with an initial condition of one exposed person at time zero and *R*_0_ *=* 3.18. Figure 1 shows the death rate, required ward and intensive care beds, numbers of susceptible, exposed, and infectious, and cumulative deaths over time. Of note is the great speed at which the disease spreads, with almost all deaths occurring within 70 days. Secondly, is the fact that at the peak of the wave, the capacity of the heath service, even assuming it was exclusively available to Covid-19 patients, is exceeded by a factor of around 50 and 4 for intensive care and ward beds respectively.

**Table 1.** Summary model results for unmitigated spread.

| Cumulative Deaths | Final Susceptible Proportion | Peak death rate (Day) | Peak Ward beds (Day) | Peak ICU beds (Day) |
| --- | --- | --- | --- | --- |
| 638,000 | 4.8% | 18,000 (106) | 638,000 (102) | 232,000 (109) |

**Figure 1.**
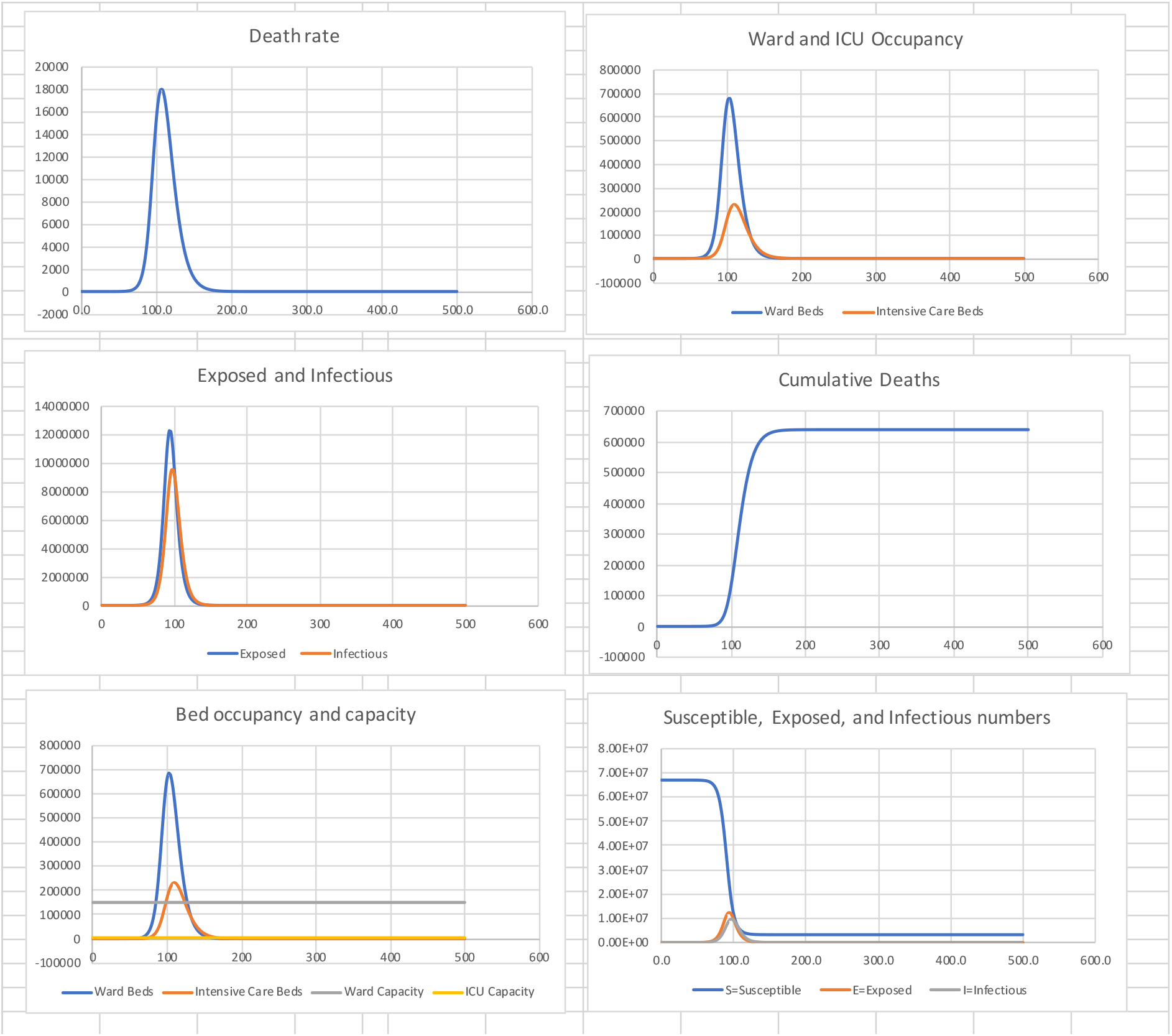
Unmitigated Spread for UK.

## 7. An example suppression

Next I consider the effect of a lockdown such as the one implemented on 23 March 2020. I use death data for the period 15 to 30 April to estimate *λ* = −0.054, with 95% confidence interval (−0.058, −0.050) and an explanatory *R*^2^ of 0.96. Thus, the estimated half-life in declining the peak, once the effect of the previous *R*_0_ of 3.18 has been largely eliminated, is 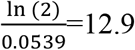 days. Using (18) this gives a reproduction number of *R*_0_ = 0.60. It is relevant to note that this is quite close to the value of 0.62 arrived at by Jarvis et al. (2020) from a behavioural survey of people’s distancing following lockdown. Table 2 shows summary statistics and figure 2 the associated graphs.

**Table 2:** *R*_0_ = 3.18 in (0, 72), *R*_0_ =0.60 in (72, 500)

| Cumulative Deaths | Final Susceptible Proportion | Peak death rate (Day) | Peak Ward beds (Day) | Peak ICU beds (Day) |
| --- | --- | --- | --- | --- |
| 39,000 | 94 % | 930 (88) | 36,000 (82) | 12,000 (92) |

Of note is the speed at which peak death rate is reached, some 16 days after starting suppression.

**Figure 2.**
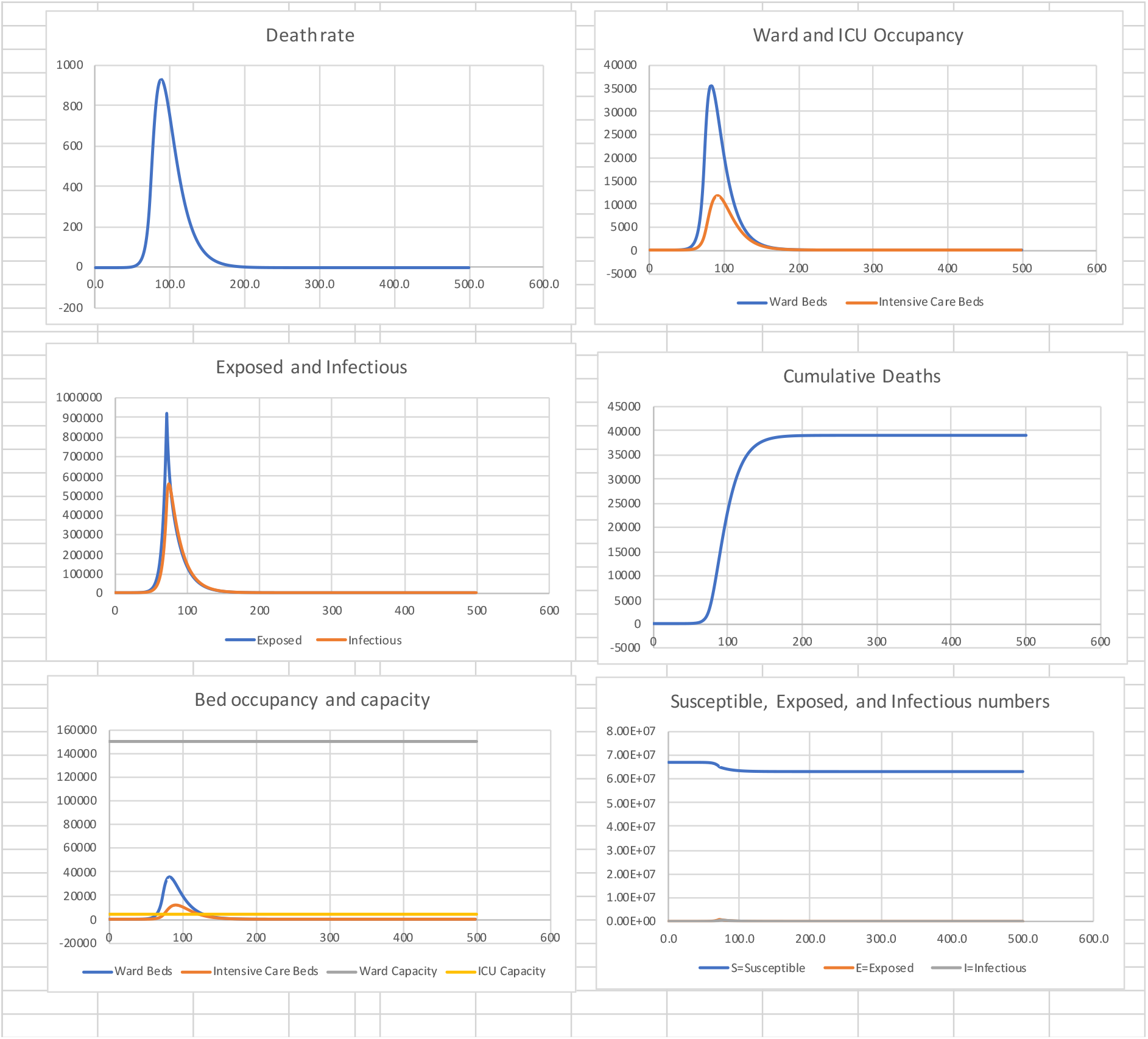
Suppression to *R*_0_ = 0.60 on day 72.

We note that ICU capacity is exceeded by a factor of 3, while ward bed demand is approximately 25% of capacity.

## 8. The effect of an earlier suppression

Now suppose that the suppression in table 2 takes place one week earlier on day 65. Table 3 and figure 3 show the model results

**Table 3:** *R*_0_ = 3.18 in (0, 65), *R*_0_ = 0.60 in (65, 500)

| Cumulative Deaths | Final Susceptible Proportion | Peak death rate (Day) | Peak Ward beds (Day) | Peak ICU beds (Day) |
| --- | --- | --- | --- | --- |
| 11,200 | 98 % | 260 (81) | 9,950 (74) | 3,350 (84) |

We see a very large reduction in absolute death numbers from around 39,000 to 11,000, This extreme sensitivity is a result of the exponential rise in infectious numbers and in hindsight clearly illustrates that earlier action was needed and would have saved many lives. The peak death rate occurs 16 days after the beginning of suppression.

**Figure 3:**
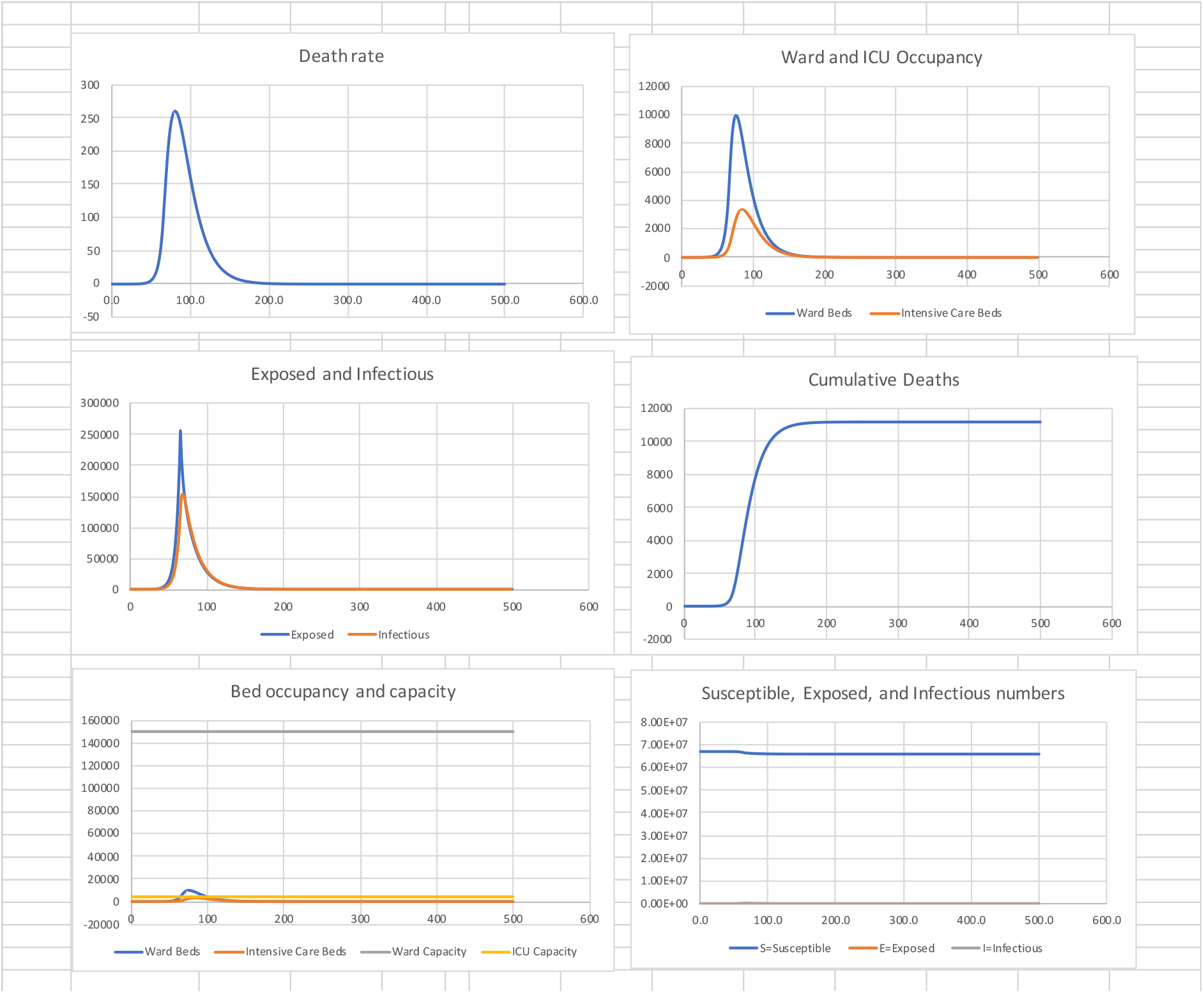
Suppression starts one week earlier.

## 9. Releasing the suppression to a relaxed *R*_0_ of 1.5

A lockdown to an *R*_0_ of 0.60 from day 72 to day 500 is unrealistic as it causes great economic and societal damage as well as increasing other mortality and morbidity. Now suppose that the suppression of figure 2 is released to a more relaxed *R*_0_ of 1.5 between days 200 and 300. That will mean there is a second wave so we limit its effect by suppressing again to *R*_0_ of 0.60 between days 300 and 500. The results are shown in table 4 and figure 4.

**Table 4:** *R*_0_ = 3.18 in (0, 72), *R*_0_ =0.60 in (72, 200), *R*_0_ =1.5 in (200, 300), *R*_0_ =0.60 in (300, 500)

| Cumulative Deaths | Final Susceptible Proportion | Peak death rate (Day) | Peak Ward beds (Day) | Peak ICU beds (Day) |
| --- | --- | --- | --- | --- |
| 42,800 | 94 % | 950 (88) | 36,500 (82) | 12,200 (91) |

**Figure 4.**
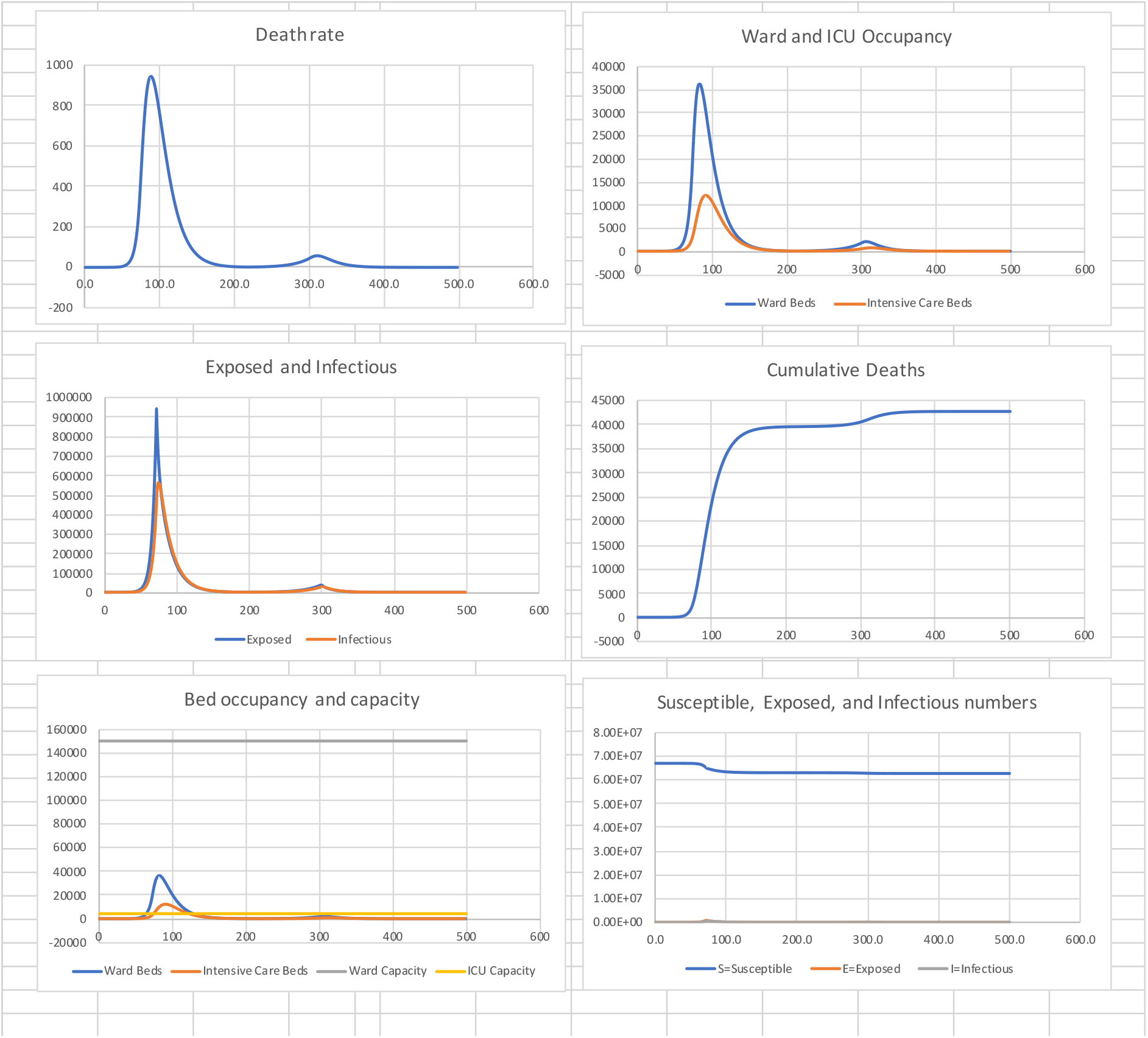
Release and second suppression.

The total deaths have been limited to around 43,000 by driving down infectious numbers to very low levels in the first wave. The first suppression was released on day 200.

## 10. Effect of early release

What happens if the release from the first suppression is brought forward from day 200 to day 180? Table 5 and figure 5 show the results.

**Table 5:**
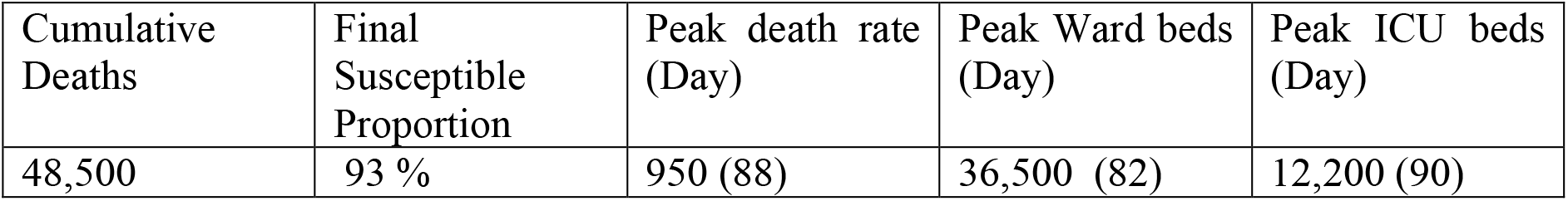
*R*_0_ = 3.18 in (0, 72), *R*_0_ =0.60 in (72, 180), *R*_0_ =1.5 in (180, 300), *R*_0_ =0.60 in (300, 500)

**Figure 5:**
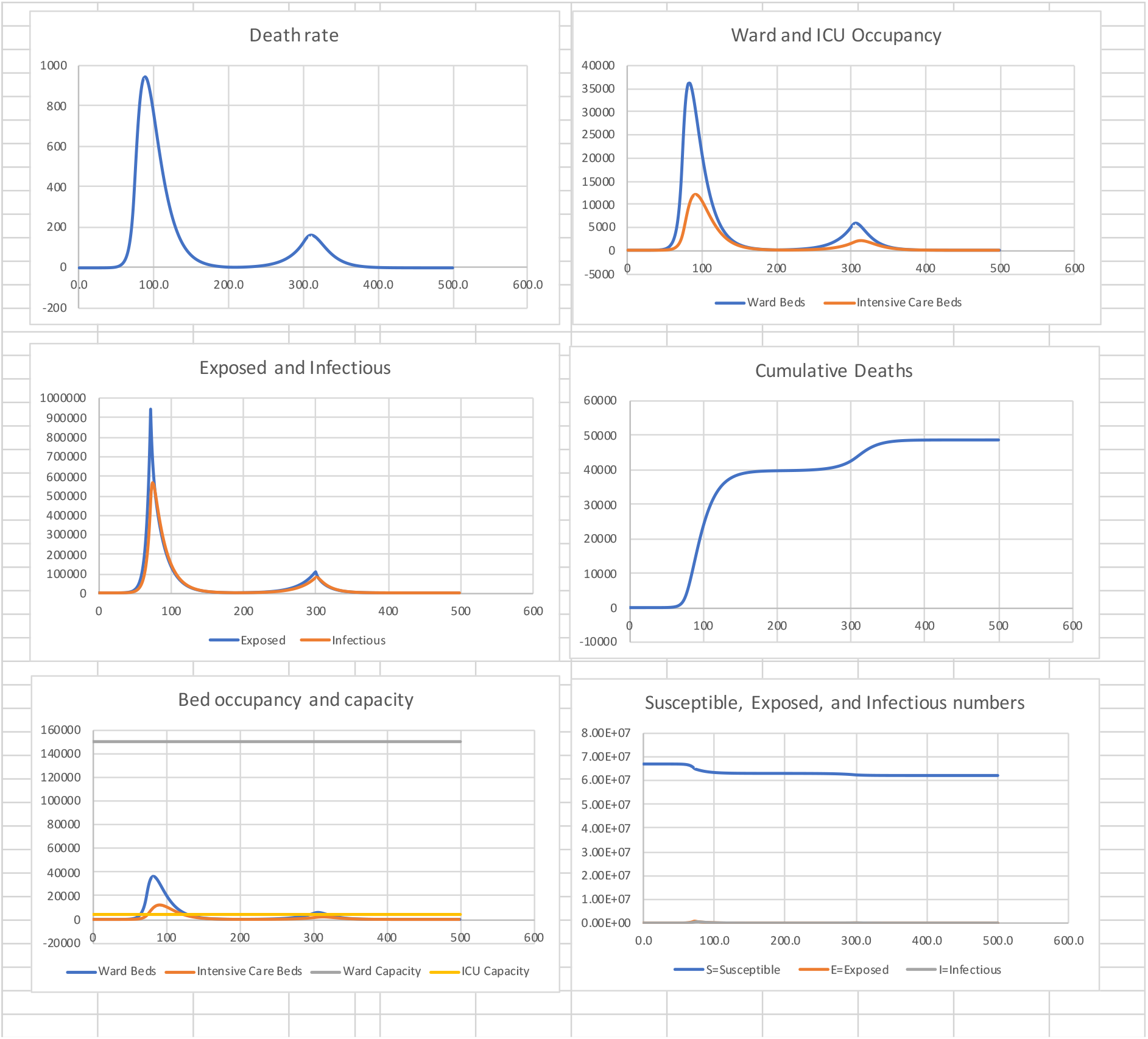
The effect of early release.

The number of deaths has now increased by around 20,000 an increase of approximately 50%.

## 11. Conclusions

The paper is motivated by a perception in the UK that the Government may have gone into lockdown too late. A literature search revealed no objective assessment as to whether this was true, and if it was, the number of lives that might have been saved had earlier action been taken. The model used here is a simple one but sufficient to give broad answers to this question. Rather than precise prediction, the intention has been to show the modelling principles and to demonstrate the extreme sensitivity of the total number of deaths to the timing of suppression measures and their relaxation. Literally, each day’s delay in starting suppression (lockdown) can result in thousands of extra deaths. The same is true for premature relaxation, acknowledging that the rate of decline is less than the rate of growth, so the effect although severe is not quite as strong. These conclusions are the incontrovertible consequence of the exponential growth and decline of a managed epidemic. As the UK attempts to manage the relaxation of measures from this first wave, a more sophisticated model might inform strategies that are spatially and temporally heterogeneous and combine shielding and segmentation of the population. These can then be put alongside the economic and other negative effects of prolonged suppression. The political decisions can then be well informed.

At the time of writing on 17 May some 55 days after the lockdown of 23 March, the total deaths were 34,000 and rising, which is perhaps quite close to the scenario considered in figure 2. Comparing with figure 3 it does pose the question as to why lockdown did not occur earlier?

Although the primary aim of the paper has been comparison of strategies rather than prediction of absolute numbers of deaths, it is noted that the cumulative deaths obtained from the model may be an underestimate. This is because a substantial proportion of deaths occurring in care homes and community are lagging those in hospitals and were not used in the estimate of *R*_0_. Further, no account has been taken in the model of a gradual transition from the unmitigated *R*_0_ of 3.18 to the estimated value of 0.60 following lockdown. A disadvantage of using death data is that one is essentially estimating *R*_0_ some 20 days previously. But the alternative of using positive test numbers might lead to biased estimates due to the volatility of the proportion of infectious persons who are represented in such figures. This was a particular problem in the UK due to the limited test capacity, particularly in the early stages. For modelling, perhaps the compromise is to use hospital admissions as a proxy for infectious numbers.

## Data Availability

Calculations available on request to author

## Footnotes

1 If ρ is the case fatality rate then the expected number of deaths up to time *t* is ρ[1 − *S*(*t − ψ)*] where *ψ* is the mean time between exposure and death. Differentiating, the death rate at 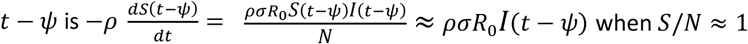 when *S*/*N* ≈ 1.

2 The Government told people who showed symptoms to stay at home, without testing for seven days before contemplating hospital admission.

3 https://www.england.nhs.uk/statistics/statistical-work-areas/covid-19-daily-deaths/

## References

Anderson, R. M., Heesterbeek, H., Klinkenberg, D., Hollingsworth, T. D. (2020), How will country based mitigation measures influence the course of the COVID-19 epidemic?, The Lancet, 21 March, 2020, vol. 395, pp931–934. https://www.thelancet.com/journals/lancet/article/PIIS0140-6736(20)30567-5/fulltext

Blackwood J. C. and Childs, L. M. (2018), An introduction to compartmental modeling for the budding infectious disease modeler, Letters in Biomathematics, 5:1, 195–221, DOI: 10.1080/23737867.2018.1509026. https://doi.org/10.1080/23737867.2018.1509026

Davies, N., Kucharski, A., Eggo, R., Gimma, A., CMMID COVID-19 working group, Edmunds, J. (2020), The effect of non-pharmaceutical interventions on COVID-19 cases, deaths and demand for hospital services in the UK: a modelling study. medRxiv preprint doi: https://doi.org/10.1101/2020.04.01.20049908

Ferguson, N. M. et al. (2020), Impact of Non-pharmaceutical interventions (NPIs) to reduce COVID-19 mortality and healthcare demand. Imperial College Response Team, Report 9, 16 March 2019, WHO Collaborating Centre for Infectious Disease Modelling, MRC Centre for Global Infectious Disease Analysis, Abdul Latif Jameel Institute for Disease and Emergency Analytics, Imperial College, London. https://www.imperial.ac.uk/mrc-global-infectious-disease-analysis/covid-19/report-9-impact-of-npis-on-covid-19/

Hethcote, H.W. (2000), The mathematics of infectious diseases, Siam Review, 42, 4, 599–653. http://links.jstor.org/sici?sici=0036-1445%28200012%2942%3A4%3C599%3ATMOID%3E2.0.CO%3B2-Q

Jarvis, C.I. et al. (2020), Quantifying the impact of physical distance measures on the transmission of COVID-19 in the UK. Centre for Mathematical Modelling of Infectious diseases, Department of Infectious diseases, London School of Hygiene and Tropical Medicine, London WC1E 7HT. First online 31 March 2020. https://www.medrxiv.org/content/10.1101/2020.03.31.20049023v1

Ma, J. (2020), Estimating epidemic exponential growth rate and basic reproduction number, Infectious disease Modelling, 5, pp 129–141 https://www.sciencedirect.com/science/article/pii/S24680427193004917.

